# Health workers’ psychological distress during early phase of the covid-19 pandemic in Morocco

**DOI:** 10.1101/2021.02.02.21250639

**Authors:** Jihane Belayachi, Sarah Benammi, Hasnae Chippo, Rhita Nechba Bennis, Naoufel Madani, Abdelmalek Hrora, Redouane Abouqal

## Abstract

**Introduction:** The mental being of healthcare workers with the COVID 19 pandemic is a determinant of their resilience. We investigated the psychological impact of health workers during early phase of Covid 19 pandemic in Morocco.

**Methods:** This was a cross-sectional study based on a survey of health workers of the Rabat University Hospital Ibn-Sina in Morocco. Data were collected during the first week of health emergency state –between 23 and 30 march-related to the covid-19 pandemic declaration in Morocco. Sociodemographic, health characteristics and professional characteristics of each health worker were collected. We also evaluated the knowledge of health workers concerning the protective measures against COVID 19. The mental health status of the health workers was investigated using the Arabic validated version of HADS 14 items, evaluating hospital anxiety and depression

**Results:** Two hundred eighty-seven health workers were included.The mean age was 34.4±12.18 year; and 64.5% were female, 54% have been trained regarding protection procedures, and 94.8% declared that they are aware of individual protection measures. The incidence of anxiety and depression was respectively 77.4% and 73.9%. High degree of anxiety and depression was associated with female gender. However, Higher degree of anxiety was also related to function, specialty of practice, and knowledge of the protective measures against COVID-19.

**Conclusion:** We reported the result of the first evaluation of psychological burden of health worker during early period of COVID-19 pandemic in a developing country. The study showed high frequency of anxiety and depression among Moroccan health worker in a hospital faced to COVID-19 patient management.

## INTRODUCTION

Since its emergence in Wuhan China, the novel Coronavirus Disease 2019 (Sars Cov 2) has become a nationwide health crisis, indeed classified by the World Health Organization (WHO) as a pandemic since early March 2020(1). To date 27 April 2020, there have been over 2,99 M confirmed cases of COVID-19 globally, and 207 k deaths worldwide. Amongst reported deaths over 100 healthcare professionals decease have been registered (2), in Morocco we report overall 4.065 confirmed cases, and 161deaths. With the outbreak of the COVID-19 in China in December 2019 and in Europe in February 2020, the adaptive capacity of a health system appears to be more depending on the relevance and precaution of the preventive measures implemented than on maturity of health care system. On March 19, health emergency state was declared in Morocco.

Moroccan authorities deployed necessary resource to fight COVID-19. Collectives preventives measures had started including; containment of population, and barrier measures (social distancing and mask wearing in public places) expanded screening of contact; and isolation of suspected and confirmed cases).

The increasing number of overwhelmed healthcare systems, reported global shortage of protective equipment, and clinical care challenges during these times, create a state of uncertainty regarding the situation progression among population in general and health care workers in particular. All these factors build up to create a high-risk environment to the physical and mental being of healthcare workers. Healthcare workers are the key resources of a health system. The mental being of healthcare workers with the COVID 19 pandemic is a determinant of their resilience. Indeed, we suppose that our global current context may become a source of psychological distress upon healthcare workers ready to face COVID 19 pandemic. Therefore, we sought to investigate the psychological burden of healthcare Workers during the early phase of the COVID-19 pandemic times.

## Methods

### Study Design and Setting

This was a cross-sectional study based on a survey of healthcare workers of the Rabat University Hospital Ibn-Sina in Morocco.

Data were collected between 23th to 30th March 2020; the first week of health emergency state related to the covid-19 pandemic declaration in Morocco. Sociodemographic, health characteristics and professional characteristics of each health worker were collected. We also evaluated the knowledge of health workers concerning the protective measures against COVID 19 through 3 questions: a) Do you know about individual protective measures? ; b)What kind of individual protective measures do you know? ; c) did you have a training regarding protection procedures?

Data were collected using face-to-face questionnaires, and by phone calls. The study was approved by the local ethics committee and informed consent was obtained from all participants.

## INSTRUMENT

The mental health status of the health workers was investigated using the Arabic validated version of HADS 14 items, evaluating hospital anxiety and depression (3). It is a self-report rating scale of 14 items on a 4-level scale (range 0–3) evaluating anxiety and depression (7 items for each subscale). The total score is the sum of the 14 items, and for each subscale the score is the sum of the respective seven items (ranging from 0–21). Scores of 8 or more on either subscale are considered to be a significant ‘case’ of psychological morbidity, and 0–7 are considered ‘normal’.

### Statistical analysis

Continuous variables are presented as mean ± standard deviation for variables with a normal distribution, and as median and interquartile range (IQR) for variables with skewed distributions. The normality of the distribution was tested by the Kolmogorov-Smirnov test with Lilliefors correction. For categorical variables, the percentages of patients in each category were calculated. Anxiety or depression (defined as a subscale score >7) was the dependent variable. The univariate analysis was carried out using the students’ t-test for the comparison of two groups. Analysis of variance for comparison of more than two groups and Pearson correlation for the link between two quantitative variables. A two-tailed *P* value < 0.05 was considered statistically significant. Statistical analyses were carried out using SPSS for Windows (SPSS, Inc., Chicago, IL, USA).

## RESULTS

Sociodemographic and health characteristics of 287 included health workers were described in table 1. The incidence of anxiety and depression was respectively 77.4% and 73.9%. Comparison of anxiety and depression intensity according health worker’s characteristics; and kknowledge and training about individual protective measures against COVID-19 were presented in table 1. Figure 1 showed Mean anxiety scale of health workers according to protective procedures’ knowledge.

**Table 1:** Characteristics of Health workers and Comparison of anxiety and depression intensity according health care worker’s characteristics

|  | N (%) | ANXIETY<br>n = 222 (77.4%) |  | DEPRESSION<br>N=212 (73.9%) |  |
| --- | --- | --- | --- | --- | --- |
|  |  | Mean± sd or (r) | P | Mean± sd or (r) | P |
| **Age Mean (± SD)** | 34.4±12.18 | -0.05 | 0.41 | 0,06 | 0.2 |
| **Gender** |  |  | <0.001 |  | 0.001 |
| Female | 185(64.5%) | 11.56 ±5. |  | 9.46±3,9 |  |
| Male | 102(35.5%) | 8.83 ±4.5 |  | 7.93±3,7 | 0.20 |
| **Marital Status** |  |  |  |  |  |
| Married | 126(43.9%) | 10.74 ±5.4 | 0.65 | 9.25 ±3.8 |  |
| Single | 161(56.1%) | 10.47 ±4.7 |  | 8.65 ±3.9 |  |
| **Years of Practice Median [IQR]** | 4[1-16] | -0.007 | 0.91 | 0.03 | 0.5 |
| **Comorbidities** |  |  | 0.42 |  | 0.87 |
| None | 136 (47.4%) | 10.84 ±5.2 |  | 8.88 ±3.9 |  |
| Yes | 151 (52.6%) | 10.37 ±4.8 |  | 8.95 ±3.8 |  |
| **Function** |  |  | 0.04 |  | 0.45 |
| Administration | 18 ( 6.3%) | 12.43 ±4.6 |  | 8.77 ±4.3 |  |
| Nurse | 72 (25.1%) | 11.36 ±4.8 |  | 9.65 ±3.7 |  |
| Health Technician | 50 (17.4) | 10.81 ±5.4 |  | 8.75 ±3.6 |  |
| Junior DOCTOR | 117 (40.8%) | 10.26 ±4.7 |  | 8.55 ±3.9 |  |
| Senior Doctor | 30 (10.6%) | 8.54 ±5.2 |  | 8.92 ±4.2 |  |
| **Departments** |  |  | 0.001 |  | 0.48 |
| Emergency units | 49 (17.1%) | 12.48±4.8 |  | 9.00 ±3.5 |  |
| Interventional units | 31 (10.8%) | 11.70 ±4.4 |  | 9.53 ±2.9 |  |
| Laboratory units | 20 (7.1%) | 10.80 ±5.6 |  | 9.96 ±4.6 |  |
| Surgical departments | 60 (21.3%) | 10.76 ± 4.8 |  | 8.99 ±3.5 |  |
| Medical departments | 65 (23%) | 9.72 ±5 .3 |  | 8.95 ±4.4 |  |
| No Care departments | 17 (6%) | 9.53 ±5.4 |  | 8.20 ±4.2 |  |
| Intensive Care units | 31 (10.8%) | 9.38 ±3.3 |  | 7.61 ±3.9 |  |
| Acute Care unit | 9 (3.2%) | 5.22 ±2.7 |  | 8.15 ±4.9 |  |
| **Training regarding protection procedures** |  |  | 0.7 |  | 0.21 |
| Yes | 155 (54%) | 10.67 ±4.8 |  | 8.65 ±3.7 |  |
| No | 132 (46%) | 10.49 ±5.2 |  | 9.23 ±4.1 |  |
| **Knowledge about protection procedures** |  |  | 0.03 |  | 0.27 |
| No | 15 (5.2%) | 13.25 ±5.3 |  | 9.98 ±3.6 |  |
| Yes | 272 (94.8%) | 10.44 ±4.9 |  | 8.86 ±3.9 |  |
| **Kind of individual protective measures** |  |  |  |  |  |
| Hand sanitizing | 236 (82.2%) |  |  |  |  |
| Mask | 214 (74.6%) |  |  |  |  |
| Social distancing | 170 (59.2%) |  |  |  |  |
| Eye Goggles | 109 (38%) |  |  |  |  |
| Hair Net | 124 (43.2%) |  |  |  |  |
| Gown | 144 (50.2%) |  |  |  |  |
| Gloves | 130 (45.3%) |  |  |  |  |
| No Surface Touching | 112 (39%) |  |  |  |  |
| Isolation | 107 (37.3%) |  |  |  |  |
| Shoe protection | 4 (1.4%) |  |  |  |  |

**Figure:**
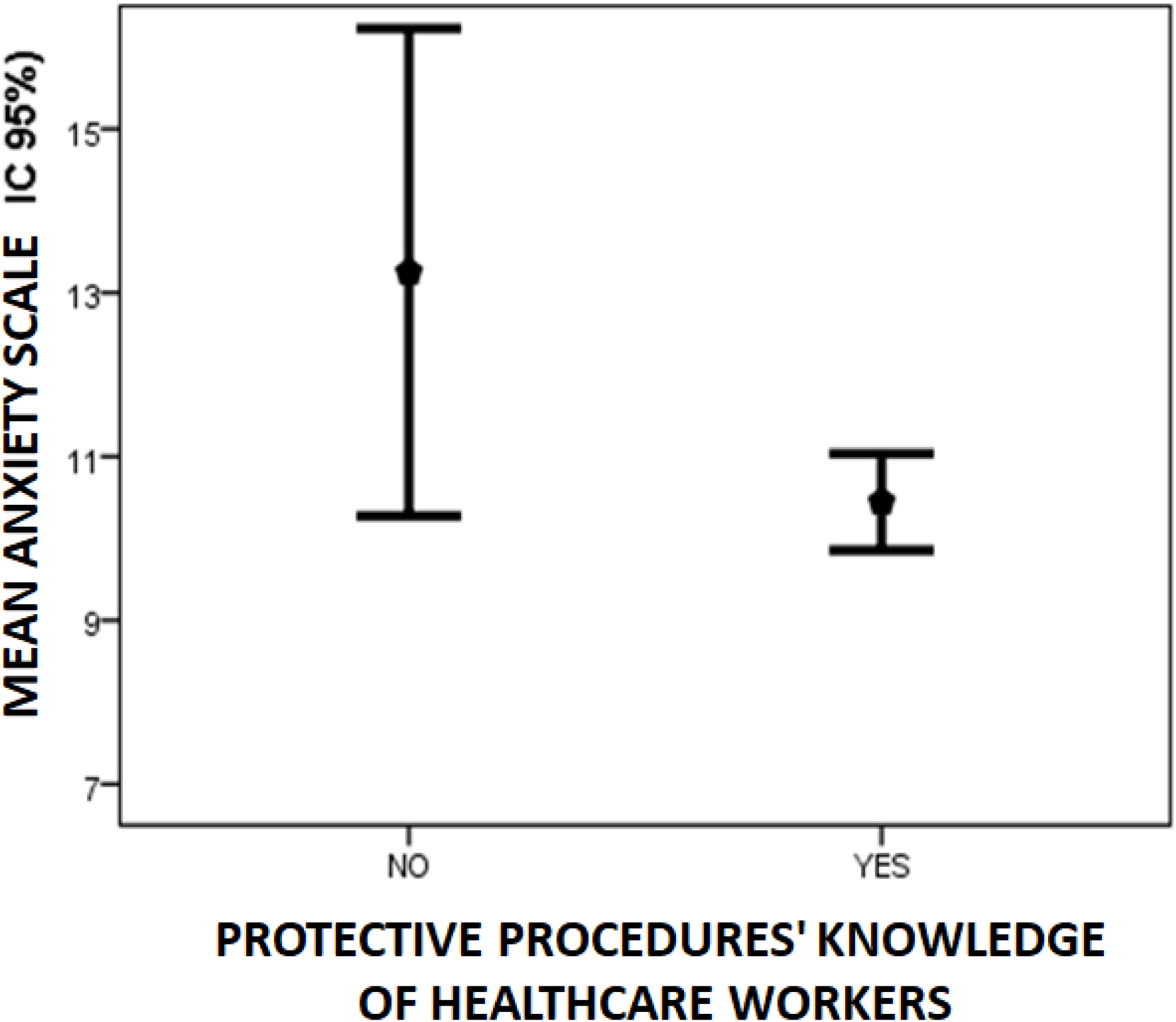
anxiety level accord to health workers Function.

## DISCUSSION

We reported the result of the first evaluation of psychological burden of health worker during early period of COVID-19 pandemic Africa. The study showed high frequency of anxiety (77.4%) and depression (73.9%) among Moroccan health worker in a hospital faced to COVID-19 patient management. High degree of anxiety and depression was associated with female gender. A previous study in a tertiary infectious disease hospital for COVID-19 in China, and concluded that female medical staff members were more vulnerable with high anxiety index (8).

However, High degree of anxiety was also related to function, specialty of practice, and knowledge of the protective measures against COVID-19. Higher anxiety was noted among emergency unit staff members, interventional units, laboratory units and surgical units. These results can be explained by the high risk of exposure proper to each department: direct manipulation of blood and high proximity to sharp instruments for surgical, laboratory and interventional units, and to frontline contact with all arriving patients to the hospital with unknown COVID-19 status regarding emergency units.

Lower knowledge of the protective measures against COVID-19 is the main factors explaining higher level of anxiety among health workers during early phase of Covid 19 pandemic. These results go in accordance with previous studies evaluating anxiety and depression among medical staff demonstrating both high anxiety and depression incidence (8).

## Data Availability

all data are available in manuscript

## Acknowledgments

none to declare

## FIGURE LEDEND

**Figure 1:** Mean anxiety scale of health workers according to protective procedures’ knowledge.

**Figure 2:** Mean anxiety level of health workers according to function

## Ethical Consideration & Disclosure

Study protocol was approved by Rabat ethic committee, and informed consent was obtained from all participants

